# Inference of SARS-CoV-2 generation times using UK household data

**DOI:** 10.1101/2021.05.27.21257936

**Authors:** W.S. Hart, S. Abbott, A. Endo, J. Hellewell, E. Miller, N. Andrews, P.K. Maini, S. Funk, R.N. Thompson

## Abstract

The distribution of the generation time (the interval between individuals becoming infected and passing on the virus) characterises changes in the transmission risk during SARS-CoV-2 infections. Inferring the generation time distribution is essential to plan and assess public health measures. We previously developed a mechanistic approach for estimating the generation time, which provided an improved fit to SARS-CoV-2 data from January-March 2020 compared to existing models. However, few estimates of the generation time exist based on data from later in the pandemic. Here, using data from a household study conducted from March-November 2020 in the UK, we provide updated estimates of the generation time. We consider both a commonly used approach in which the transmission risk is assumed to be independent of when symptoms develop, and our mechanistic model in which transmission and symptoms are linked explicitly. Assuming independent transmission and symptoms, we estimated a mean generation time (4.2 days, 95% CrI 3.3-5.3 days) similar to previous estimates from other countries, but with a higher standard deviation (4.9 days, 3.0-8.3 days). Using our mechanistic approach, we estimated a longer mean generation time (6.0 days, 5.2-7.0 days) and a similar standard deviation (4.9 days, 4.0-6.3 days). Both models suggest a shorter mean generation time in September-November 2020 compared to earlier months. Since the SARS-CoV-2 generation time appears to be changing, continued data collection and analysis is necessary to inform future public health policy decisions.

## INTRODUCTION

The generation time of a SARS-CoV-2 infector-infectee pair is defined as the period of time between the infector and infectee each becoming infected [1–5]. The distribution of the generation times of many infector-infectee pairs characterises the temporal profile of infectiousness of an infected host (averaged over all hosts and normalised so that it represents a valid probability distribution) [6]. Inferring the generation time distribution of SARS-CoV-2 is important in order to predict the effects of non-pharmaceutical interventions such as contact tracing and quarantine [7, 8]. In addition, the generation time distribution is widely used in epidemiological models for estimating the reproduction number from case notification data [6, 9–11] and is crucial for understanding the relationship between the reproduction number and the epidemic growth rate [3, 6].

The SARS-CoV-2 generation time distribution has previously been estimated using data from known infector-infectee transmission pairs [8, 12, 13] or entire clusters of cases [14–16]. These studies involved data [8, 14, 17–20] collected between December 2019 and April 2020, almost all from countries in East and Southeast Asia (with the exception of four transmission pairs from Germany and four from Italy in [8]). Evidence from January and February 2020 in China suggested a temporal reduction in the mean generation time due to non-pharmaceutical interventions [15]. Specifically, effective isolation of infected individuals is likely to have reduced the proportion of transmissions occurring when potential infectors were in the later stages of infection, thereby shortening the generation time [15]. Similarly, two other studies found a decrease in the serial interval (the difference between symptom onset times of an infector and infectee) [21] and an increase in the proportion of presymptomatic transmissions [22] in China over the same time period, which can be attributed to symptomatic hosts being isolated increasingly quickly over time.

Despite estimation of the SARS-CoV-2 generation time in Asia early in the pandemic, relatively little is known about the generation time distribution outside Asia, and whether or not any changes have occurred in the generation time since the early months of the pandemic. In particular, we are aware of only one study in which the generation time was estimated using data from the UK [23]. In that study [23], data describing symptom onset dates for 50 infector-infectee pairs, collected by Public Health England between January and March 2020 as part of the “First Few Hundred” case protocol [24, 25], were used to infer the generation time distribution. However, since these transmission pairs mostly consisted of international travellers and their household contacts, the authors concluded that their estimates of the generation time may have been biased downwards due to enhanced surveillance and isolation of these cases [23].

Here, we use data from a household study, conducted between March and November 2020, to estimate the SARS-CoV-2 generation time distribution in the UK under two different underlying transmission models. In the first model (the “independent transmission and symptoms model”), a parsimonious assumption is made that the generation time and the incubation period of the infector are independent (i.e., there is no link between the times at which infectors transmit the virus and the times at which they develop symptoms), as has often been employed in studies in which the SARS-CoV-2 generation time has been estimated [8, 12–14, 23, 26] (Table 1). In the second model (the “mechanistic model”), we use a mechanistic approach in which potential infectors progress through different stages of infection, first becoming infectious before developing symptoms [12]. Infectiousness is therefore explicitly linked to symptoms in the mechanistic model. By fitting separately to data from three different time intervals within the study period, we explore whether or not there was a detectable temporal change in the generation time distribution. Our study represents the first analysis of the SARS-CoV-2 generation time outside Asia conducted using data from after the earliest stages of the pandemic.

**Table 1.**
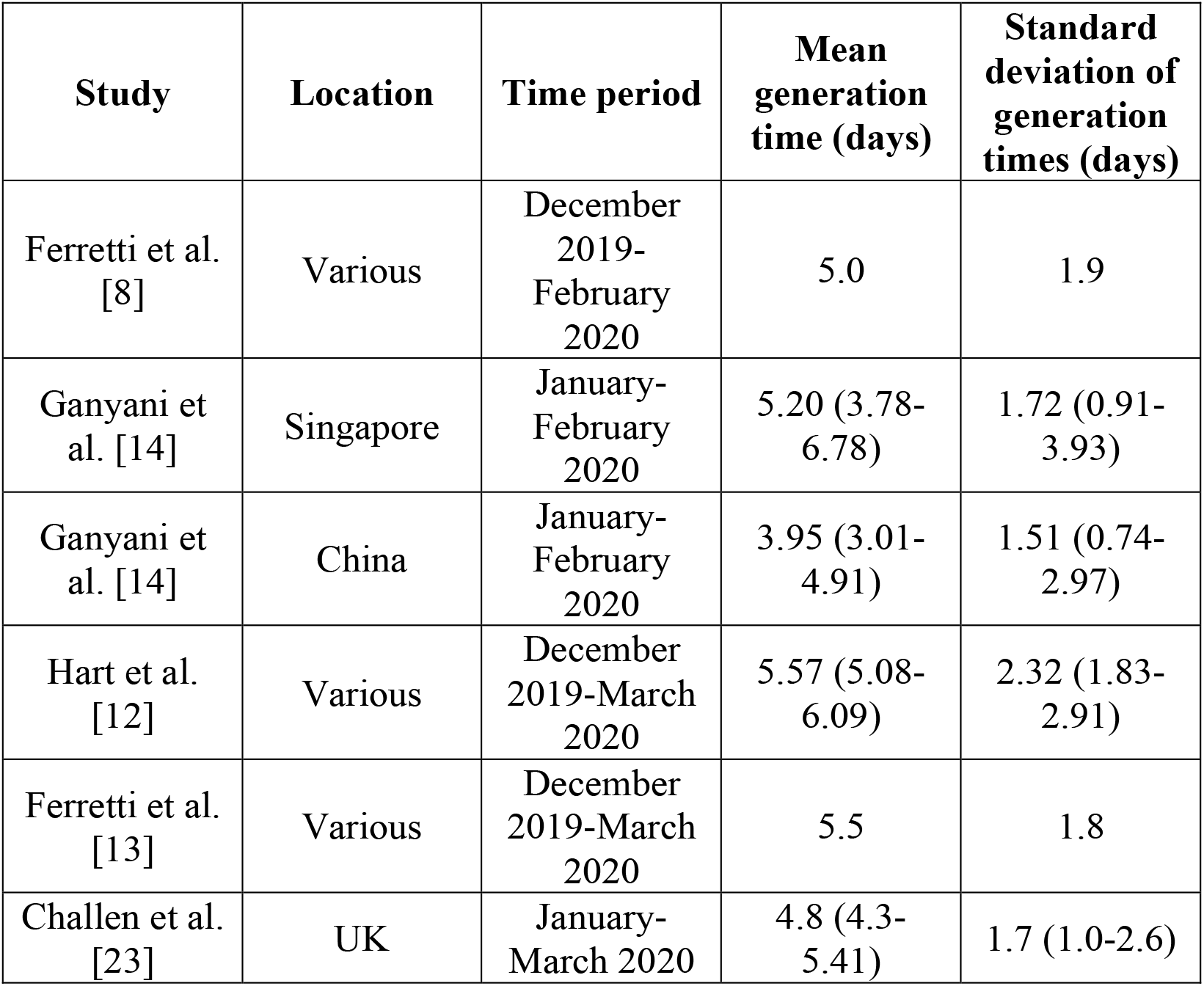
Previous SARS-CoV-2 generation time estimates. Estimates of the mean and standard deviation of the generation time distribution, obtained under the assumption of independent transmission and symptoms. 95% credible intervals are shown in brackets where available.

| Study | Location | Time period | Mean generation time (days) | Standard deviation of generation times (days) |
| --- | --- | --- | --- | --- |
| Ferretti et al. [8] | Various | December 2019-February 2020 | 5.0 | 1.9 |
| Ganyani et al. [14] | Singapore | January-February 2020 | 5.20 (3.78-6.78) | 1.72 (0.91-3.93) |
| Ganyani et al. [14] | China | January-February 2020 | 3.95 (3.01-4.91) | 1.51 (0.74-2.97) |
| Hart et al. [12] | Various | December 2019-March 2020 | 5.57 (5.08-6.09) | 2.32 (1.83-2.91) |
| Ferretti et al. [13] | Various | December 2019-March 2020 | 5.5 | 1.8 |
| Challen et al. [23] | UK | January-March 2020 | 4.8 (4.3-5.41) | 1.7 (1.0-2.6) |

## RESULTS

### Inferring generation times from UK household data

We used data augmentation MCMC to fit two models of infectiousness (the independent transmission and symptoms model and the mechanistic model) to data describing individuals’ infection status and symptom onset dates in 172 UK households (see Methods). For the two fitted models, we calculated posterior estimates of the mean (Figure 1A) and standard deviation of the generation time distribution (Figure 1B), in addition to the proportion of transmissions occurring prior to symptom onset (among infectors who develop symptoms; Figure 1C) and the overall infectiousness parameter, *β*_0_ (see Methods; Figure 1D). Under the commonly used independent transmission and symptoms model, we obtained a point estimate (posterior mean) of 4.2 days (95% CrI 3.3-5.3 days) for the mean generation time (Figure 1A, blue violin). This value is similar to a previous estimate using data from China in [14], and is slightly lower than estimates for Singapore in [14] and for several countries (predominantly in Asia) in [8] (Table 1), although those estimates lie within our credible interval. On the other hand, our estimated standard deviation of 4.9 days (95% CrI 3.0-8.3 days; Figure 1B, blue violin) is substantially higher than previous estimates (Table 1). Using our mechanistic model, we obtained a higher estimate for the mean generation time of 6.0 days (95% CrI 5.2-7.0 days; Figure 1A, red violin), and a similar estimate for the standard deviation (4.9 days, 95% CrI 4.0-6.3 days; Figure 1B, red violin), compared to those predicted by the independent transmission and symptoms model. The two models gave similar posterior distributions for the proportion of transmissions prior to symptom onset (Figure 1C), with point estimate values of model parameters generating estimates of 0.72 (95% CrI 0.63-0.80) for the independent transmission and symptoms model, and 0.73 (95% CrI 0.61-0.83) for the mechanistic model.

**Figure 1.**
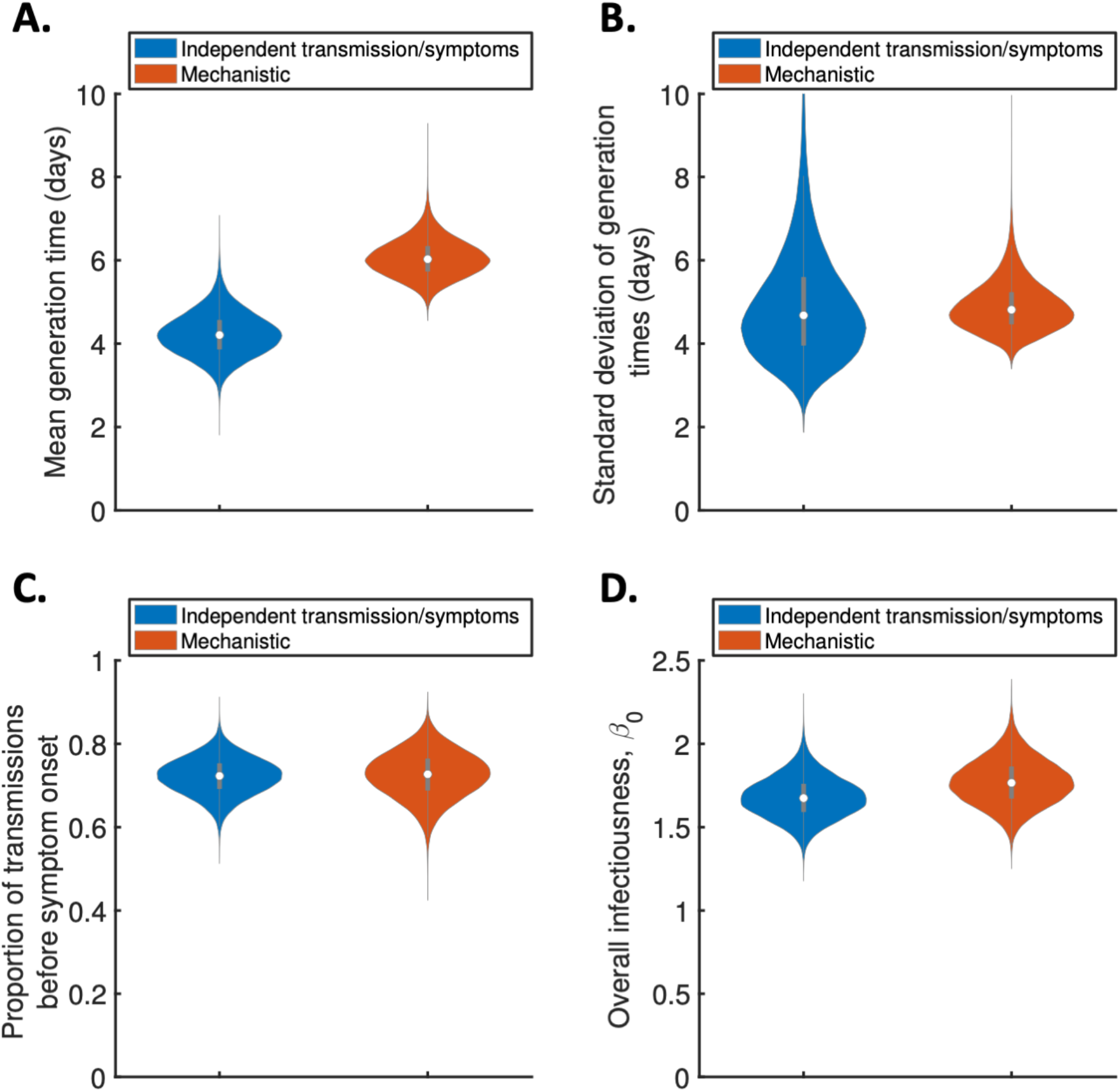
Comparison of posterior predictions. Violin plots indicating posterior distributions of the mean generation time (A), standard deviation of generation times (B), proportion of transmissions occurring prior to symptom onset (among infectors who develop symptoms; C), and overall infectiousness parameter, *β*_0_ (D). We show results obtained both using a model in which infectiousness is assumed to be independent of when symptoms develop (“independent transmission and symptoms model”, blue), and using the mechanistic approach from [12] in which infectiousness is explicitly linked to symptoms (“mechanistic model”, red).

Posterior distributions and point estimates (posterior means) for all fitted model parameters are shown in the Supplementary Material (Figures S1-S2 and Tables S2-S3). Since only the likelihood with respect to augmented data was calculated in the MCMC procedure, explicit comparisons of goodness of fit between the models were not readily available. However, comparing model predictions of the difference between successive symptom onset dates with the UK household data indicated that both models provided a similar fit to the data (Figure S3).

As described in Methods, the generation time distribution that we considered in Figure 1 (and elsewhere, unless otherwise stated) corresponds to the normalised expected infectiousness profile of a host at each time since infection. However, realised generation times are expected to be shorter for infected individuals in small households compared to those in large households. This is due to depletion of susceptible household members in small households before longer generation times can be attained [6, 27]. As a result, we also predicted the mean and standard deviation of realised generation times within the study households (Figure S4A-B), accounting for the precise distribution of household sizes in the study. For both the independent transmission and symptoms model and the mechanistic model, point estimates for the mean (3.6 days and 4.9 days for the two models, respectively) and standard deviation (3.8 days and 4.1 days) of realised household generation times were lower than those shown in Figure 1. Since transmission then typically occurred earlier in the infector’s course of infection than shown in Figure 1, we predicted a higher proportion of presymptomatic transmissions within the study households (Figure S4C) compared to the estimates in Figure 1C.

For both models, we then used point estimates of fitted model parameters to infer the distributions of the generation time (Figure 2A), the time from onset of symptoms to transmission (TOST; Figure 2B) and the serial interval (Figure 2C). The TOST distribution (which characterises the relative expected infectiousness of a host at each time from symptom onset, as opposed to from infection [13, 20, 26, 28, 29]) predicted using the mechanistic model was more concentrated around the time of symptom onset compared to that obtained using the independent transmission and symptoms model (Figure 2B), as was found in [12]. In contrast, the estimated serial interval distributions were similar for the two models (Figure 2C).

**Figure 2.**
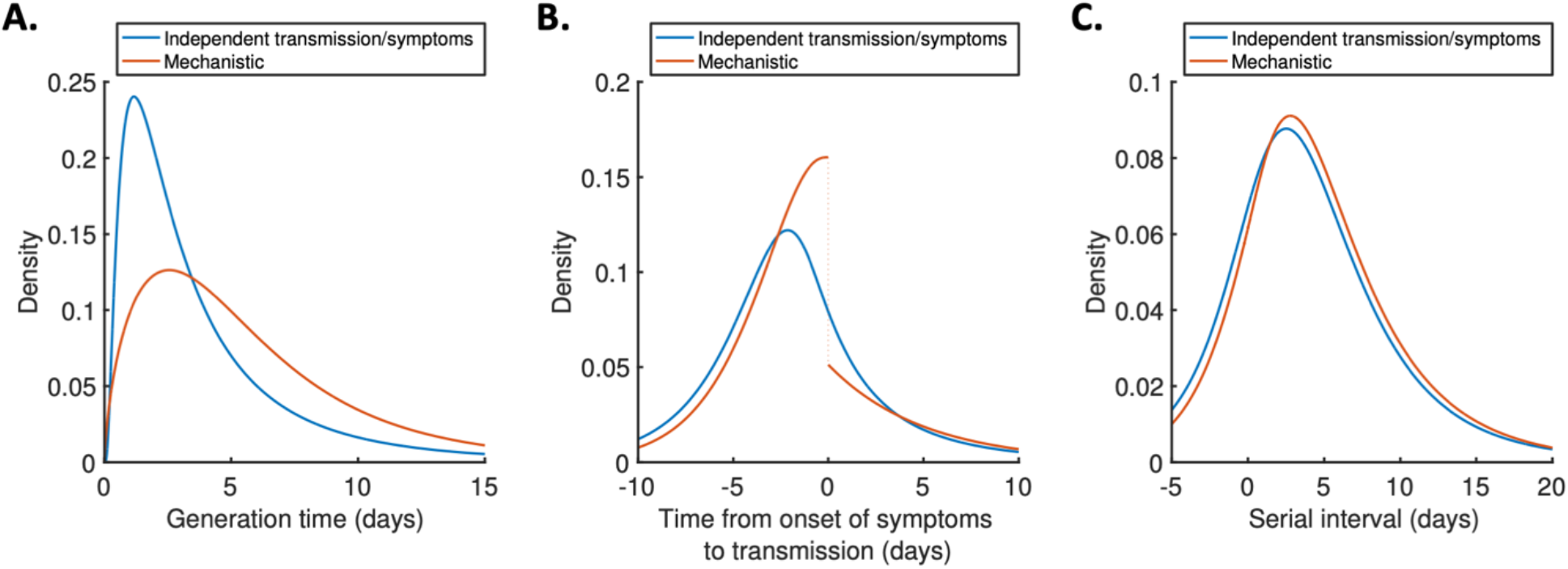
Generation time, TOST and serial interval distributions. Inferred generation time (A), TOST (B) and serial interval (C) distributions for the two models, obtained using point estimate (posterior mean) parameters. Note that the discontinuity in the red curve in Figure 2B occurs because two different transmission rates were fitted for infectors in the presymptomatic and symptomatic periods. The reduction in transmission following symptom onset can be attributed to changes in behaviour in response to symptoms [30].

### Temporal variation in the generation time distribution

To explore whether or not the generation time distribution changed during the study period, we separately fitted the independent transmission and symptoms model to the data from households in which the index case was recruited in (i) March-April, (ii) May-August, or (iii) September-November 2020 (Figure 3). We chose these time periods to ensure the numbers of households recruited into the study during each interval were similar (Figure S5).

**Figure 3.**
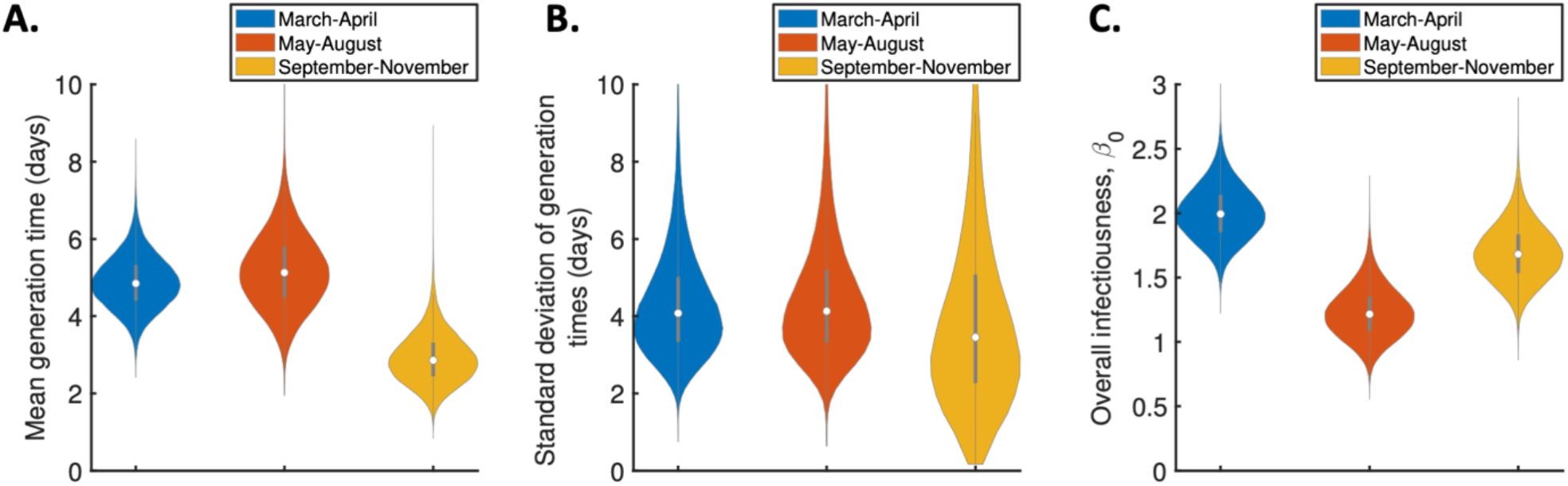
Temporal changes in the generation time distribution. Violin plots indicating posterior distributions of the mean generation time (A), standard deviation of generation times (B), and overall infectiousness parameter, *β*_0_ (C), for the independent transmission and symptoms model fitted to data from March-April (blue), May-August (red) or September-November 2020 (orange).

The results shown in Figure 3A suggest a lower mean generation time in September-November (2.9 days, 95% CrI 1.8-4.3 days) compared to earlier months (4.9 days, 95% CrI 3.6-6.3 days, for March-April and 5.2 days, 95% CrI 3.4-7.2 days, for May-August). A similar temporal reduction in the mean generation time was found when we instead fitted the mechanistic model to the data from the three time intervals (Figure S6A). To explore the lower estimated generation time for September-November further, we also fitted the independent transmission and symptoms model to the data from each of these months individually (Figure S7). The shorter estimated generation time compared to earlier in the pandemic was consistent in each of the three months (Figure S7A).

### Sensitivity analyses

In the independent transmission and symptoms model, we assumed that both the generation time and incubation period followed lognormal distributions. The mean and standard deviation of the generation time distribution were estimated by fitting the model to the household transmission data. In the fitting procedure, we assumed that the incubation period followed a lognormal distribution that was obtained in a previous meta-analysis [31]. In contrast, we assumed in our mechanistic approach that each infection could be decomposed into three gamma distributed stages (latent, presymptomatic infectious and symptomatic infectious), so that the incubation period was also gamma distributed (with the same mean and standard deviation as the lognormal distribution obtained in [31]). An expression for the generation time distribution in the mechanistic model, which did not take a simple parametric form, is given in the Supplementary Text. However, we conducted supplementary analyses in which we instead assumed that either the generation time (Figure S8A-C) or incubation period (Figure S8D-F) in the independent transmission and symptoms model was gamma distributed. In both cases, we obtained similar results to those shown for that model in Figure 1.

In Figure S9, we relaxed the assumption of a fixed incubation period distribution, using the confidence intervals obtained in [31] to account for uncertainty in the incubation period distribution (Figure S9A-B). For both the independent transmission and symptoms model and the mechanistic model, accounting for this uncertainty did not substantially affect posterior estimates of either the mean (Figure S9C) or the standard deviation (Figure S9D) of the generation time distribution.

In our main analyses, we assumed that household transmission was frequency-dependent, so that the force of infection exerted by an infected household member on each susceptible household member scaled with 1/*n*, where *n* is the household size. In order to explore the robustness of our results to this assumption, we considered alternative possibilities where infectiousness scaled with *n*^−*ρ*^, for *ρ* = 0 (density-dependent transmission) and *ρ* = 0.5 (Figure S10A-B). We also conducted an analysis in which the dependency, *ρ*, was estimated alongside other model parameters (Figure S10C). We found that our estimates of the mean and standard deviation of the generation time distribution were robust to the assumed value of *ρ* (Figure S10A-B). However, when the value *ρ* was fitted (Figure S10C), we estimated a value of 1.0 (95% CrI 0.6-1.5). This supported our assumption of frequency-dependent transmission, although the credible interval was relatively wide. In addition, we considered the possibility that infectiousness instead scales with 1/(*n* − 1), so that the infectious individual is removed from the scaling in the force of infection *β*(*τ*), and again obtained similar estimates of the mean and standard deviation of the generation time distribution compared to those shown in Figure 1 (Figure S10D-E).

We also considered the sensitivity of our results to the assumed relative infectiousness of asymptomatic infected hosts (Figure S11). In most of our analyses, we assumed that the expected infectiousness of an infected host who remained asymptomatic throughout infection was a factor *α*_*A*_ = 0.35 times that of a host who develops symptoms, at each time since infection [31]. However, similar estimates of the mean (Figure S11A) and standard deviation (Figure S11B) of the generation time distribution were obtained when we instead assumed *α*_*A*_ = 0.1 or *α*_*A*_ = 1.27 (these values corresponded to the lower and upper confidence bounds obtained in [32]).

Finally, we explored the robustness of our results to the exclusion of hosts with unknown infection status (see Methods), considering the extreme possibilities where these hosts were instead assumed to have either all remained uninfected, or all become infected (Figure S12). Although the estimates of *β*_0_ were affected by this assumption (Figure S12C), the estimated generation time distribution was robust to the assumed infection status of these hosts (Figure S12A-B).

## DISCUSSION

In this study, we estimated the generation time distribution of SARS-CoV-2 in the UK by fitting two different models to data describing the infection status and symptom onset dates of individuals in 172 households. The first of these models was predicated on an assumption that transmission and symptoms are independent. While this assumption has often been made in previous studies in which the SARS-CoV-2 generation time has been estimated [8, 14, 23, 33, 34], it is not an accurate reflection of the underlying epidemiology [26, 35]. Therefore, we also considered a mechanistic model based on compartmental modelling, which was shown in [12] to provide an improved fit to data from 191 SARS-CoV-2 infector-infectee pairs compared to previously used models that have been used to estimate the generation time. Here, infection times and the order of transmissions within households were unknown, whereas in [12] the direction of transmission was assumed to be known for each infector-infectee pair. For that reason, we used data augmentation MCMC to fit the two models to the household data, in a similar fashion to previous studies of household influenza virus transmission [27, 36, 37].

Under the model assuming independent transmission and symptoms, we estimated a mean generation time of 4.2 days (95% CrI 3.3-5.3 days) and a standard deviation of 4.9 days (95% CrI 3.0-8.3 days). The estimate of the mean generation time was comparable to previous estimates obtained under this assumption using data from elsewhere [8, 13, 14] (Table 1). On the other hand, while our credible interval for the standard deviation was wide, the estimates obtained in those previous studies [8, 13, 14] all lay below our lower 95% credible limit of 3.0 days. One potential cause of this disparity is the difference in isolation policies for symptomatic hosts between countries. In particular, the UK’s policy of self-isolation may be expected to lead to a longer-tailed generation time distribution compared to countries with a policy of isolation outside the home, since under home isolation, some within-household transmission is likely to occur even following isolation. Isolation outside the home was commonplace in the East and Southeast Asian countries where the majority of the data underlying the estimates in [8, 13, 14] were collected. Because we used only household transmission data, this effect was likely to be particularly pronounced in our estimates.

Using the mechanistic model, we predicted a higher mean generation time of 6.0 days (95% CrI 5.2-7.0 days) compared to the value estimated under the assumption of independent transmission and symptoms. On the other hand, the inferred serial intervals for the independent transmission and symptoms model and mechanistic model were similar (Figure 2C), with means of 4.2 days and 4.7 days, respectively. Temporal information in our household transmission data consisted mostly of symptom onset dates, with very few hosts testing positive before developing symptoms. Therefore, the variation in estimates of the generation time between the models can be attributed to differences in the assumed relationships between the generation time and serial interval under those models. For the independent transmission and symptoms model, the generation time and serial interval distributions have the same mean, as is commonly assumed to be the case [26]. However, this was not true for the mechanistic model, in which hosts with longer presymptomatic infectious periods generate (on average) a higher number of transmissions. As a result, under the mechanistic model, a randomly chosen infection is likely to arise from an infector with a longer than expected incubation period, thereby leading to a longer generation time than serial interval (an analytical expression for the exact difference between the mean generation time and serial interval for that model is derived in the Supplementary Material).

Our results did not indicate any clear difference in goodness of fit between the two models (Figure S3). A range of factors should therefore be considered when deciding which of our estimates of epidemiological parameters to use in subsequent analyses. Although any model requires simplifying assumptions to be made, our mechanistic approach allows the standard assumption of independent transmission and symptoms to be relaxed by providing a mechanistic underpinning to the relationship between the times at which individuals display symptoms and become infectious. Furthermore, as described above, this model was shown in [12] to provide a better fit to another SARS-CoV-2 dataset than a model assuming independence between transmission and symptoms (in [12], the simpler setting of transmission pairs rather than households facilitated direct model comparison). On the other hand, the independent transmission and symptoms model has the advantage of producing an estimated generation time distribution with a simple parametric form. The choice of estimates to use may also depend on precisely what the estimates are being used for. For example, the generation time distribution inferred under the assumption of independent transmission and symptoms may be better suited for use in some models for estimating the time-dependent reproduction number, since those models often also implicitly involve the assumption that transmission and symptoms are independent [10]. In contrast, the parameter estimates from our mechanistic approach correspond naturally to parameters in compartmental epidemic models.

By fitting separately to data from three different intervals within the study period (March-November 2020), we investigated whether or not the generation time distribution in the UK changed as the pandemic progressed. Our results indicate a lower mean generation time in September-November compared to earlier months (Figure 3A). One possible explanation for this is a higher proportion of time spent indoors in colder months leading to an increased household transmission risk, particularly in the early stages of infection before symptoms develop (since symptomatic infected hosts are still likely to self-isolate). While shorter generation times may in general be over-represented in data collected from infector-infectee pairs at times when case numbers are rising [8, 26, 38] (as was the case in September and October [39, 40]), we estimated the mean generation time to be similar in November (when national case numbers were mostly decreasing [39, 40]) compared to September and October (Figure S7).

Our finding of a temporal decrease in the mean generation time during the study period highlights the importance of obtaining up-to-date generation time estimates specific to the location under study. Should variations in the generation time distribution occur and not be accounted for, estimates of the time-dependent reproduction number may be incorrect [3, 41]. Specifically, if the mean generation time is shorter than assumed, then the true value of the time-dependent reproduction number is closer to one than the inferred value [3] (and vice versa).

The B1.1.7 SARS-CoV-2 variant [42], which has since become the dominant lineage in the UK, was responsible for infections in only two households within our dataset. Therefore, additional data are needed to quantify the impact of the emergence of this variant on the generation time. A shorter mean generation time of the B1.1.7 variant has been hypothesised as one possible contribution to the increased growth rate of cases in the UK in late 2020 and early 2021, but a decrease in the generation time alone was not found to explain trends in UK case data [43, 44]. Conversely, viral load data suggest a longer duration of viral shedding due to infection with B1.1.7 compared to the original variant of SARS-CoV-2 [45]. If higher viral loads lead to increased infectiousness [46–50], this may suggest a longer-tailed generation time distribution for the B1.1.7 variant.

One advantage of our approach is that we are able to include the contribution of asymptomatic infected hosts to household transmission chains in our analyses. We showed that our estimated generation time distribution was robust to the assumed relative infectiousness of infected hosts who remained asymptomatic, *α*_*A*_ (Figure S11). Similarly, while we assumed frequency-dependent household transmission in most of our analyses, we found that the exact relationship between the household size and transmission rate had little effect on our estimates of the mean and standard deviation of the generation time distribution (Figure S10). We also considered estimating the exponent governing the dependency of transmission on household size (Figure S10C). This supported our assumption of frequency-dependent transmission. In previous studies of influenza transmission within households, evidence has been found both in favour of [36] and against [51] frequency-dependent transmission.

Our study has some limitations. Since we used household transmission data in our analyses, the generation time for transmission outside the household may differ from our estimates. Future extensions to our approach may account for the possibility that more than one household member was infected in the same primary infection event or the potential for multiple sequential introductions of the virus into a household [37]. Allowing for multiple introductions may shorten estimates of the generation time, although any effect will dependent significantly on the community prevalence and the number of contacts that household members have with individuals in the community. In contrast, accounting for potential co-primary infections is likely to lead to higher estimates of the generation time.

Other further work may include exploring heterogeneity in the generation time distribution between individuals and/or households with different characteristics. This could involve, but is not limited to, estimating the generation time distribution for individuals of different age, sex and ethnicity.

In summary, we have inferred the SARS-CoV-2 generation time distribution in the UK using household data and two different transmission models. A key output of this research is one of the only estimates of the SARS-CoV-2 generation time outside Asia. Another crucial feature of our analysis is that it was based on data from beyond the first few months of the pandemic. Since this research suggests that the generation time may be changing, continued data collection and analysis is of clear importance.

## METHODS

### Data

Data were obtained from a household study conducted by Public Health England in 172 households (with 603 household members in total) between March and November 2020. In each household, an index case was recruited following a positive PCR test. The following were then recorded for each household member:

- The timing and outcome of (up to) two subsequent PCR tests.
- The outcome of an antibody test (carried out for 90% of individuals).
- Whether or not the household member developed symptoms.
- The date of symptom onset (only for symptomatic individuals with a positive PCR or antibody test).

In the study, all household members who tested positive in either a PCR or antibody test were assumed to have been infected. Conversely, all individuals who tested negative for antibodies (and where the two PCR tests were either negative or were not carried out) were assumed to have remained uninfected, irrespective of symptom status. For 6% of the study cohort, no antibody test was carried out and any PCR tests were negative. These hosts with unknown infection status were excluded from our main analyses (but were counted in the household size), although we also considered the sensitivity of our results to this assumption.

In two households, at least one household member developed symptoms 55-56 days prior to the symptom onset date of the index case, with no other household members developing symptoms (or returning a positive PCR or antibody test) between these dates. In contrast, the maximum gap between successive symptom onset dates in the remaining households was 25 days. Data from these two households were excluded from our analyses, on the basis that the virus was most likely introduced multiple times into these households.

### Models

#### General modelling framework

We denote the force of infection exerted by a given infected host onto each susceptible member of their household, at time *τ* days since infection, by *β*(*τ*), where we assumed

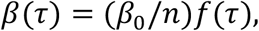

if the host develops symptoms, and

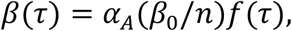

if the host remains asymptomatic. Here:

- *β*_0_ represents the overall infectiousness of a host who develops symptoms.
- *n* is the household size. The scaling of *β*(*τ*) with 1/*n* corresponds to frequency-dependent transmission, as assumed in [36, 52], although we considered alternative possibilities where transmission was either density-dependent (without the scaling factor 1/*n*) or scaled with 1/*n*^0.5^ [51] in sensitivity analyses.
- *f*(*τ*) is the generation time distribution.
- *α*_*A*_ is the relative infectiousness of infected hosts who remain asymptomatic. We assumed a value of 0.35 [32] in most of our analyses, although we also carried out sensitivity analyses in which different values of *α*_*A*_ were considered.

Throughout, the normalised infectiousness profile, *f*(*τ*), is referred to as the generation time distribution. However, realised generation times within a household may be shortened due to the depletion of susceptible household members before longer generation times can be attained [6, 27]. For example, if an infected host was equally infectious at two times since infection, *τ*_1_ < *τ*_2_, then we would have *f*(*τ*_1_) = *f*(*τ*_2_). However, because the number of susceptible household members may decrease between these two times (i.e., either the host under consideration, or another infected household member, may transmit the virus within the household in the intervening time), then household transmission is in fact more likely to occur in the household at the earlier time, *τ*_1_, when more susceptibles are available. Therefore, we also predicted the mean and standard deviation of realised generation times within the study households in the Supplementary Material (Figure S4).

We considered two different models of infectiousness, which are outlined below. Under each model, expressions were derived in [12] for the generation time, time from onset of symptoms to transmission (TOST) and serial interval distributions, in addition to the proportion of transmissions occurring before symptom onset. These expressions are given in the Supplementary Material (other than the generation time distribution and proportion of presymptomatic transmissions for the independent transmission and symptoms model, which are stated below).

#### Independent transmission and symptoms model

In this model, the infectiousness of an infected host (who does not remain asymptomatic throughout infection; asymptomatic infected hosts are considered separately) at a given time since infection, *τ* days, is assumed to be independent of whether or not the host has yet developed symptoms – i.e., the generation time and incubation period are independent. In this case, the generation time distribution, *f*(*τ*), was assumed to be the probability density function of a lognormal distribution [37] (an alternative case of a gamma distributed generation time is considered in Figure S8). The mean and standard deviation of this distribution, in addition to *β*_0_, were estimated when we fitted the model to the household transmission data.

Under the assumption of independent transmission and symptoms, the proportion of transmissions occurring prior to symptom onset (among infectors who develop symptoms) is given by [8, 53]

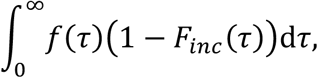

where *F*_*inc*_ is the cumulative distribution function of the incubation period (this was assumed to be known; the exact incubation period distribution we used is given under “Parameter estimation” below).

#### Mechanistic model

Under the mechanistic model [12], infectors who develop symptoms progress through independent latent (*E*), presymptomatic infectious (*P*) and symptomatic infectious (*I*) stages. The duration of each stage was assumed to be gamma distributed, and infectiousness was assumed to be constant during each stage. Under these assumptions, an explicit expression can be derived for the expected infectiousness, *β*(*τ* ∣ *τ*_*inc*_), of a host (who develops symptoms) at time since infection *τ*, conditional on their incubation period *τ*_*inc*_. Details of the mechanistic approach, including the formula for *β*(*τ* ∣ *τ*_*inc*_), are provided in the Supplementary Material.

When we fitted this model to the household transmission data, three model parameters were estimated in addition to *β*_0_. These parameters correspond to:

- The ratio between the mean latent (*E*) period and the mean incubation (combined *E* and *P*) period (where the latter was assumed known).
- The mean symptomatic infectious (*I*) period.
- The ratio between the transmission rates when potential infectors are in the *P* and *I* stages.

### Likelihood function

The models were fitted to the household data using data augmentation Markov chain Monte Carlo (MCMC; see further details below). For a household (of size *n*) in which *n*_*I*_ household members are infected during the study (of whom *n*. develop symptoms and *n*_*A*_ remain asymptomatic throughout infection) and *n*_*U*_ = *n* – *n*_*I*_ are uninfected, augmented data consist of the entire sequence of infection times of individuals in the household 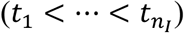, as well as the exact symptom onset times (*t*_*s,j*_) of each host, *j*, who develops symptoms.

When deriving an expression for the likelihood contribution from the household, given these augmented data, we made several simplifying assumptions:

- The virus was introduced once into each household (i.e., no subsequent infections from the community occurred following the infection of the primary case).
- No co-primary cases.
- Potential bias towards more recent infection of the primary host when community prevalence was increasing, or less recent when prevalence was decreasing [8, 26, 38], was neglected.

We denote the expected infectiousness of household member *j*, at time *τ* since infection, by *β*_*j*_(*τ*). For the mechanistic model in which transmission and symptoms are not independent, this infectiousness is conditional on the duration of the incubation period, *t*_*s,j*_ – *t*_*j*_, if host *j* developed symptoms. The total (instantaneous) force of infection exerted at time *t* on each susceptible household member is then

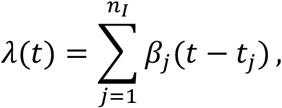

where *β*_*j*_(*t* – *t*_*j*_) = 0 for *t* ≤ *t*_*j*_, and the cumulative force of infection is

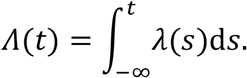

For *k* = 2, …, *n*_*I*_, conditional on the sequence of infection times up to time *t*_*k*_, the probability that host *k* becomes infected at time *t*_*k*_ is given by

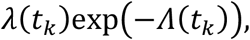

where exp(−*Λ*(*t*_*k*_)) represents the probability that host *k* avoided infection up to time *t* [36, 37]. For *k* = *n*_*I*_ + 1, …, *n*, conditional on the entire sequence of infection times, 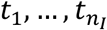, the probability that host *k* remains uninfected is given by exp(−*Λ*(∞)). In the case of independent transmission and symptoms, we have

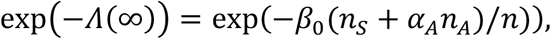

whereas for the mechanistic model, exp(−*Λ*(∞)) instead depends on the incubation periods of those hosts who develop symptoms (see the Supplementary Material).

The likelihood contribution from the household, *L*(*θ*), where *θ* is the vector of unknown model parameters, given the augmented data, can therefore be written as

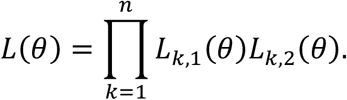

Here, *L*_*k*,1_(*θ*) is the contribution to the likelihood from the transmission, or absence of transmission, to host *k*, i.e.,

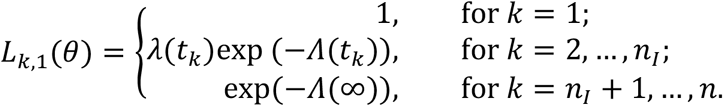

*L*_*k*,2_(*θ*) is the contribution from the incubation period of host *k* (where applicable), i.e.,

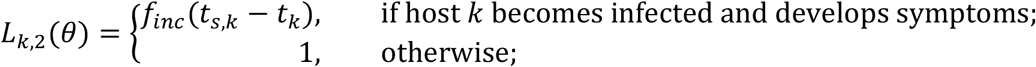

where *f*_*inc*_ is the probability density function of the incubation period (this was assumed to be known; the exact incubation period distribution we used is given below).

Each household was assumed to be independent, so that the overall likelihood was given by the product of the contributions from each household.

### Parameter estimation

#### Incubation period

For the independent transmission and symptoms model, we assumed a lognormal incubation period distribution with mean 5.8 days and standard deviation 3.1 days [31]. For the mechanistic model, we assumed a gamma distributed incubation period with the same mean and standard deviation; this was for mathematical convenience, since the incubation period was decomposed into the sum of independent gamma distributed latent and presymptomatic infectious periods. Results for the independent transmission and symptoms model using a gamma distributed incubation period are shown in Figure S8, and we account for uncertainty in the exact parameters of the incubation period distribution in Figure S9.

#### Parameter fitting procedure

Unknown model parameters were estimated using data augmentation MCMC. The observed data comprised information about whether or not individuals were ever infected and/or displayed symptoms, symptom onset dates, and for some individuals an upper bound on the infection time (corresponding to the date of a positive PCR test). These data were augmented with precise times of infection and symptom onset (where applicable) for each host. No prior assumptions were made about the order of transmissions within the household.

Below, we outline the parameter fitting procedure that we used for the independent transmission and symptoms model. The procedure used for the mechanistic model was similar and is described in the Supplementary Material.

Lognormal priors were assumed for fitted model parameters (these parameters were the mean and standard deviation of the generation time distribution, in addition to the overall infectiousness, *β*_0_). The priors for the mean and standard deviation of the generation time distribution had medians of 5 days and 2 days, respectively (these choices were informed by previous estimates of the SARS-CoV-2 generation time distribution [8, 13, 14]), and were chosen to ensure a prior probability of only 0.025 that these parameters exceeded very high values of 10 days and 7 days, respectively. The exact priors we used are detailed in Table S2.

We denote the vector of model parameters by *θ*, and the augmented data by

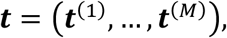

where ***t***^(*m*)^ represents the augmented data from household *m* = 1, …, *M*, and *M* is the total number of households. We write the (overall) likelihood as

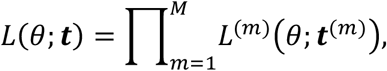

where the likelihood contribution, *L*^(*m*)^(*θ*; ***t***^(*m*)^), from each household, *m*, was computed as described in the previous section, and denote the prior density of *θ* by *π*(*θ*).

In each step of the chain, we carried out (in turn) one of the following:

1. Propose new values for the vector of model parameters, *θ*, using independent normal proposal distributions for each parameter (around the corresponding parameter values in the previous step of the chain). Accept the proposed parameters, *θ*_*prop*_, with probability

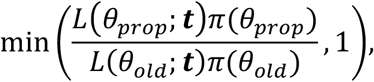

where *θ*_*old*_ denotes the vector of parameter values from the previous step of the chain, and where the augmented data, ***t***, remain unchanged in this step.
2. Propose new values for the precise symptom onset times of each symptomatic infected host, using independent uniform proposal distributions (within the day of symptom of onset for each host). For each household, *m*, accept the proposed augmented data, 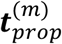, from that household with probability

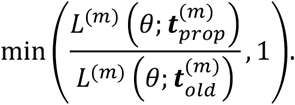

where 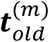 denotes the corresponding augmented data from the previous step of the chain, and where the model parameters, *θ*, remain unchanged in this step (i.e., proposed times are accepted/rejected independently for each household, according to the likelihood contribution from that household).
3. Propose new values for the infection time of one randomly chosen symptomatic infected host in each household (in households where there was at least one), using independent normal proposal distributions (around the equivalent times in the previous step of the chain). For each household, *m*, accept the proposed augmented data, 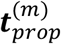, from that household with probability

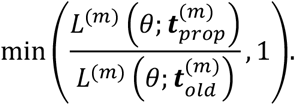
4. Propose new values for the infection time of one randomly chosen asymptomatic infected host in each household (in households where there was at least one), using independent normal proposal distributions (around the equivalent times in the previous step of the chain). For each household, *m*, accept the proposed augmented data, 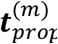, from that household with probability

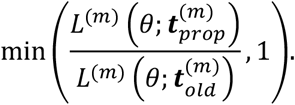

The chain was run for 10,000,000 iterations; the first 2,000,000 iterations were discarded as burn-in. Posteriors were obtained by recording only every 100 iterations of the chain.

## Governance statement

The household study was approved by the PHE Research Ethics and Governance Group as part of the portfolio of PHE’s enhanced surveillance activities in response to the pandemic.

## Data Availability

Data and code will be made publicly available upon publication, but are available upon request to the authors.

## Acknowledgments

Thanks to Pauline Waight, who managed the data for the household study, and to the PHE staff who collected the data and tested the PCR and serum samples. Thanks also to Rob Challen, Julia Gog, Matt Keeling and other members of the Juniper Consortium (www.maths.org/juniper/) for helpful comments about this research.

