## Supplementary Material for "Inference of SARS-CoV-2 generation times using UK household data"

**Supplementary Text**:

<u>Details of mechanistic model</u>

Each infected host (who developed symptoms) was assumed to progress through latent (*E*), presymptomatic infectious (*P*) and symptomatic infectious (*I*) stages. We denote the infectiousness of hosts in the *P* and *I* stages by $\beta_P$ and $\beta_I$, respectively, and define the ratio $\alpha_P = \beta_P/\beta_I$. The duration of each stage, denoted $y_{E/P/I}$, was assumed to be gamma distributed:

$$y_E \sim \text{Gamma}(k_E, 1/(k_{inc}\gamma)),$$

$$y_P \sim \text{Gamma}(k_P, 1/(k_{inc}\gamma)),$$

$$y_I \sim \text{Gamma}(k_I, 1/(k_I\mu)),$$

where we write $X \sim \text{Gamma}(a, b)$ for a gamma distributed random variable with shape parameter $a$ and scale parameter $b$. We assumed that $k_E + k_P = k_{inc}$, so that the incubation period, $\tau_{inc} = y_E + y_P$, was gamma distributed, with

$$\tau_{inc} \sim \text{Gamma}(k_{inc}, 1/(k_{inc}\gamma)).$$

Here, the parameters $k_{inc}$ and $1/\gamma$ (which represent the shape parameter of the incubation period distribution and the reciprocal of the mean incubation period, respectively) were fixed in order to obtain the specified incubation period distribution (the exact values that we assumed are given in Table S1).

For simplicity, we assumed that $k_I = 1$, so the symptomatic infectious period was exponentially distributed. The parameters $k_E$ (the shape parameter of the latent (*E*) period distribution), $1/\mu$ (the mean symptomatic infectious (*I*) period) and $\alpha_P$ (the ratio between the transmission rates of hosts in the *P* and *I* stages) were estimated when we fitted the model to the household transmission data.

Hosts who remained asymptomatic throughout infection were assumed to follow the same *E*/*P*/*I* stages, although in this case the distinction between the *P* and *I* stages has no epidemiological meaning. Stage durations, as well as the value of $\alpha_P$, were assumed to be identical for entirely asymptomatic hosts and those who developed symptoms, so that the generation time distribution was the same for all hosts.

*Conditional infectiousness*

For a host who develops symptoms, conditional on incubation period $\tau_{inc}$, the expected infectiousness at time since infection $\tau$ is [1]

$$\beta(\tau \mid \tau_{inc}) = \begin{cases} \alpha_P C(\beta_0/n)(1 - F_{Beta}(1 - \tau/\tau_{inc}; k_P, k_E)), & 0 < \tau < \tau_{inc}, \\ C(\beta_0/n)(1 - F_I(\tau - \tau_{inc})), & \tau > \tau_{inc}. \end{cases}$$

Here, $\beta_0$ is the overall infectiousness parameter (see Methods in the main text), $n$ is the household size, $F_I(y)$ is the cumulative distribution of the duration of the *I* stage, $F_{Beta}(x; a, b)$ is the cumulative distribution of a beta distributed random variable with shape parameters $a$ and $b$, and

$$C = \frac{k_{inc}\gamma\mu}{\alpha_P k_P \mu + k_{inc}\gamma}.$$

The cumulative conditional infectiousness can therefore be calculated to be

$$B(\tau \mid \tau_{inc}) = \int_0^{\tau} \beta(\tilde{\tau} \mid \tau_{inc}) \mathrm{d}\tilde{\tau}$$

$$= \begin{cases} (\tau - \tau_{inc})\beta(\tau \mid \tau_{inc}) + \dfrac{\alpha_P C\beta_0}{n}\left[\dfrac{k_p\tau_{inc}}{k_{inc}}\left(1 - F_{Beta}(1 - \tau/\tau_{inc}; k_P + 1, k_E)\right)\right], & 0 \leq \tau < \tau_{inc}, \\ (\tau - \tau_{inc})\beta(\tau \mid \tau_{inc}) + \dfrac{C\beta_0}{n}\left[\dfrac{\alpha k_p\tau_{inc}}{k_{inc}} + \dfrac{1}{\mu}F_{Gamma}\left(\tau - \tau_{inc}; k_I + 1, \dfrac{1}{k_I\mu}\right)\right], & \tau \geq \tau_{inc}, \end{cases}$$

where $F_{Gamma}(x; a, b)$ is the cumulative distribution of a gamma distributed random variable with shape parameter $a$ and scale parameter $b$. The total force of infection on each household member (over the course of infection) is then

$$B(\infty \mid \tau_{inc}) = \frac{\beta_0}{n}\left(\frac{\alpha k_P \gamma \mu \tau_{inc} + k_{inc}\gamma}{\alpha k_P \mu + k_{inc}\gamma}\right).$$

The mean of this expression over the incubation period distribution is then $\beta_0/n$.

For hosts who remained asymptomatic throughout infection, conditional on the combined duration of the *E* and *P* stages, $\tau_{inc} = y_E + y_P$, the infectiousness, $\beta(\tau \mid \tau_{inc})$, was given by $\alpha_A$ times the corresponding expression for those who developed symptoms. We note that in this case, $\tau_{inc}$ has no epidemiological interpretation, but this conditional infectiousness was useful when fitting parameters (see "Parameter fitting" below).

*Generation time distribution*

The generation time, $\tau_{gen}$, can be written as

$$\tau_{gen} = y_E + y^*,$$

where $y_E$ is the length of the latent (*E*) stage, and $y^*$ is the time from the start of the presymptomatic infectious (*P*) stage to the transmission occurring. As shown in [1], $y^*$ has density

$$f^*(y^*) = C\left(\alpha_P\big(1 - F_P(y^*)\big) + \int_0^{y^*} \big(1 - F_I(y^* - y_P)\big) f_P(y_P)\mathrm{d}y_P\right).$$

Using this density, it can be shown that the moments of this distribution are

$$E[(y^*)^m] = \frac{C}{m+1}\left(\alpha_P E[{y_P}^{m+1}] + E[(y_P + y_I)^{m+1} - {y_P}^{m+1}]\right).$$

In particular, we have

$$E[y^*] = \frac{C}{2}\left(\alpha_P E[{y_P}^2] + 2E[y_P]E[y_I] + E[{y_I}^2]\right),$$

$$\mathrm{Var}[y^*] = \frac{C}{3}\left(\alpha_P E[{y_P}^3] + 3E[{y_P}^2]E[y_I] + 3E[y_P]E[{y_I}^2] + E[{y_I}^2]\right) - (E[y^*])^2.$$

Note that for a gamma distributed random variable, $X \sim \mathrm{Gamma}(a, b)$, we have

$$E[X^m] = \frac{\Gamma(a+m)}{\Gamma(a)} b^m = a(a+1)\dots\big(a+(m-1)\big)b^m.$$

Therefore, for gamma distributed stage durations, explicit expressions can be obtained for the mean and variance of the generation time distribution,

$$E[\tau_{gen}] = E[y_E] + E[y^*],$$

$$\mathrm{Var}[\tau_{gen}] = \mathrm{Var}[y_E] + \mathrm{Var}[y^*],$$

since $y_E$ and $y^*$ are assumed to be independent.

*Proportion of presymptomatic transmissions*

Among infectors who develop symptoms, the proportion of transmissions occurring prior to symptom onset is given by [1]

$$q_P = \frac{\left(\frac{\beta_P k_P}{k_{inc}\gamma}\right)}{\left(\frac{\beta_P k_P}{k_{inc}\gamma} + \frac{\beta_I}{\mu}\right)} = \frac{\alpha_P k_P \mu}{\alpha_P k_P \mu + k_{inc}\gamma}.$$

*Parameter fitting*

The vector of model parameters,

$$\theta = (k_E/k_{inc}, 1/\mu, \alpha_P, \beta_0),$$

was estimated by fitting the mechanistic model to the household transmission data.

We assumed independent prior distributions for each entry of $\theta$. Lognormal priors were assumed for $1/\mu$, $\alpha_P$ and $\beta_0$. Since $\alpha_P$ represents the ratio between the transmission rate of hosts in the *P* and *I* stages, a prior with median 0 was used to ensure equal prior probabilities of values above and below 1. This prior was also chosen to limit the prior probability of extreme values, with a prior 95% credible interval of [0.2,5]. A beta prior was used for $k_E/k_{inc}$ (which was constrained to lie between 0 and 1), and was chosen to restrict the prior probability of values very close to either 0 or 1. The exact priors we used are given in Table S3.

A slightly amended version of the parameter fitting algorithm described in the main text for the independent transmission and symptoms model was used. In particular, we augmented the observed data with:

i. The infection time, $t_j$, of each infected host.
ii. The time, $t_{s,j}$, at which each infected host transitioned from the *P* to *I* stage.

Note that for hosts who developed symptoms, the time of entry into the *I* stage corresponds to the symptom onset time. The data were also augmented with this transition time for entirely asymptomatic infected hosts because the conditional infectiousness, $\beta(\tau \mid t_{s,j} - t_j)$, was relatively easy to calculate compared to $\beta(\tau)$.

In each step of the chain, we carried out (in turn) one of the following:

1. Propose new values for the vector of model parameters, $\theta$, using a multivariate normal proposal distribution (around the value of $\theta$ in the previous step of the chain; a correlation of 0.5 was used between the proposal distributions of $k_E/k_{inc}$ and $\alpha_P$, and between those of $1/\mu$ and $\alpha_P$). Accept the proposed parameters, $\theta_{prop}$, with probability

$$\min\left(\frac{L(\theta_{prop};\boldsymbol{t})\pi(\theta_{prop})}{L(\theta_{old};\boldsymbol{t})\pi(\theta_{old})}, 1\right),$$

   where $\theta_{old}$ denotes the vector of parameter values from the previous step of the chain, and where the augmented data, $\boldsymbol{t}$**,** remain unchanged in this step.
2. Propose new values for the precise symptom onset times of each symptomatic infected host, using independent uniform proposal distributions (within the

day of symptom of onset for each host). For each household, $m$, accept the proposed augmented data, $\boldsymbol{t}_{prop}^{(m)}$, from that household with probability

$$\min\left(\frac{L^{(m)}\left(\theta; \boldsymbol{t}_{prop}^{(m)}\right)}{L^{(m)}\left(\theta; \boldsymbol{t}_{old}^{(m)}\right)}, 1\right),$$

where $\boldsymbol{t}_{old}^{(m)}$ denotes the corresponding augmented data from the previous step of the chain, and where the model parameters, $\theta$, remain unchanged in this step (i.e., proposed times are accepted/rejected independently for each household, according to the likelihood contribution from that household).

3. Propose new values for the infection time of one randomly chosen infected host in each household (either symptomatic or asymptomatic), using independent normal proposal distributions (around the equivalent times in the previous step of the chain). For each household, $m$, accept the proposed augmented data, $\boldsymbol{t}_{prop}^{(m)}$, from that household with probability

$$\min\left(\frac{L^{(m)}\left(\theta; \boldsymbol{t}_{prop}^{(m)}\right)}{L^{(m)}\left(\theta; \boldsymbol{t}_{old}^{(m)}\right)}, 1\right).$$

4. Propose new values for both the infection time, $t$, and the time of the start of the $I$ stage, $t_s$, holding $t_s - t$ constant, for one randomly chosen asymptomatic infected host in each household (in households where there was at least one), using independent normal proposal distributions (around the equivalent times in the previous step of the chain). For each household, $m$, accept the proposed augmented data, $\boldsymbol{t}_{prop}^{(m)}$, from that household with probability

$$\min\left(\frac{L^{(m)}\left(\theta; \boldsymbol{t}_{prop}^{(m)}\right)}{L^{(m)}\left(\theta; \boldsymbol{t}_{old}^{(m)}\right)}, 1\right).$$

#### Relationship between generation time, TOST and serial interval

Here, we consider a randomly chosen infector-infectee pair (in which both the infector and the infectee develop symptoms) within a large, well-mixed population. In that setting, the observed distribution of generation times is equal to the normalised infectiousness profile, which will not be true within a household (cf. Figure 1 and Figure S4). We define:

$$\tau_{inc,1} = (\text{incubation period of the infector}),$$

$$\tau_{inc,2} = (\text{incubation period of the infectee}),$$

$$\tau_{gen} = (\text{generation time}),$$

$$x_{tost} = \big(\text{time from onset of symptoms (of infector) to transmission (TOST)}\big),$$

$$x_{ser} = (\text{serial interval}),$$

where we use $\tau$ for time intervals relative to the time of infection and $x$ for those relative to the time of symptom onset. We denote the probability density functions of these time periods by $f_{inc,1}$, $f_{inc,2}$, $f_{gen}$, $f_{tost}$ and $f_{ser}$, respectively. Note that we have

$$x_{tost} = \tau_{gen} - \tau_{inc,1},$$

and

$$x_{ser} = x_{tost} + \tau_{inc,2},$$

so that

$$x_{ser} = \tau_{gen} + \tau_{inc,2} - \tau_{inc,1}.$$

In the independent transmission and symptoms model, $\tau_{gen}$ and $\tau_{inc,1}$ are assumed to be independent, and the incubation periods of the infector and infectee are assumed to be drawn independently from the population incubation period distribution, $f_{inc} = f_{inc,1} = f_{inc,2}$. Therefore, the TOST distribution is given by the convolution

$$f_{tost}(x_{tost}) = \int_0^\infty f_{gen}(x_{tost} + \tau) f_{inc}(\tau)\, \mathrm{d}\tau. \tag{1}$$

We also assume that $x_{tost}$ and $\tau_{inc,2}$ are independent, so the serial interval distribution can be calculated from the TOST distribution as

$$f_{ser}(x_{ser}) = \int_0^\infty f_{tost}(x_{ser} - \tau) f_{inc}(\tau)\, \mathrm{d}\tau. \tag{2}$$

Note that

$$E[x_{ser}] = E[\tau_{gen}] + E[\tau_{inc,2}] - E[\tau_{inc,1}] = E[\tau_{gen}],$$

i.e., the generation time and serial interval distributions have the same mean.

For the mechanistic model, we still have $f_{inc,2} = f_{inc}$, and the serial interval distribution can be calculated from the TOST distribution using equation (2). On the other hand, $\tau_{gen}$ and $\tau_{inc,1}$ are not independent, so equation (1) connecting the TOST and generation time distributions for the independent transmission and symptoms model does not hold for the mechanistic model. As shown in [1], the TOST distribution for the mechanistic model is, instead, given by

$$f_{tost}(x_{tost}) = \begin{cases} \alpha_P C(1 - F_P(-x_{tost})), & x_{tost} < 0, \\ C(1 - F_I(x_{tost})), & x_{tost} \geq 0. \end{cases}$$

Further, under the mechanistic model, the expected number of presymptomatic transmissions generated by an infected host is dependent on their incubation period. As a result, the infector's incubation period does not follow the same distribution as that of the infectee. In particular, by Bayes' theorem, we have

$$f_{inc,1}(\tau_{inc,1}) = p(\tau_{inc,1} \mid 1 \to 2) = \frac{p(1 \to 2 \mid \tau_{inc,1}) f_{inc}(\tau_{inc,1})}{p(1 \to 2)},$$

where we write $1 \to 2$ to denote the occurrence of the transmission from the infector to the infectee. Because we are here considering a large population, the probability of the transmission occurring is proportional to the overall infectiousness of the infector (integrated over the course of infection), $B(\infty)$, so we have

$$f_{inc,1}(\tau_{inc,1}) = \frac{B(\infty \mid \tau_{inc})f_{inc}(\tau_{inc,1})}{B(\infty)} = \left(\frac{\alpha_P k_P \gamma \mu \tau_{inc,1} + k_{inc}\gamma}{\alpha_P k_P \mu + k_{inc}\gamma}\right) f_{inc}(\tau_{inc,1}).$$

The expected incubation period of the infector is then

$$E[\tau_{inc,1}] = \frac{1}{\gamma} + \frac{\alpha_P k_P \mu}{k_{inc}\gamma(\alpha_P k_P \mu + k_{inc}\gamma)} = E[\tau_{inc,2}] + \frac{q_P}{k_{inc}\gamma},$$

where $q_P$ is the proportion of transmissions occurring prior to symptom onset.

As a result of the above, the expected values of the generation time and serial interval in the mechanistic model are not equal. Instead, we have

$$E[x_{ser}] = E[\tau_{gen}] - \frac{q_P}{k_{inc}\gamma}.$$

Under the values of $k_{inc}$ and $\gamma$ that we assumed (Table S1), this gives a mean generation time that is approximately $(1.6 \times q_P)$ days longer than the mean serial interval.

### Supplementary Figures:

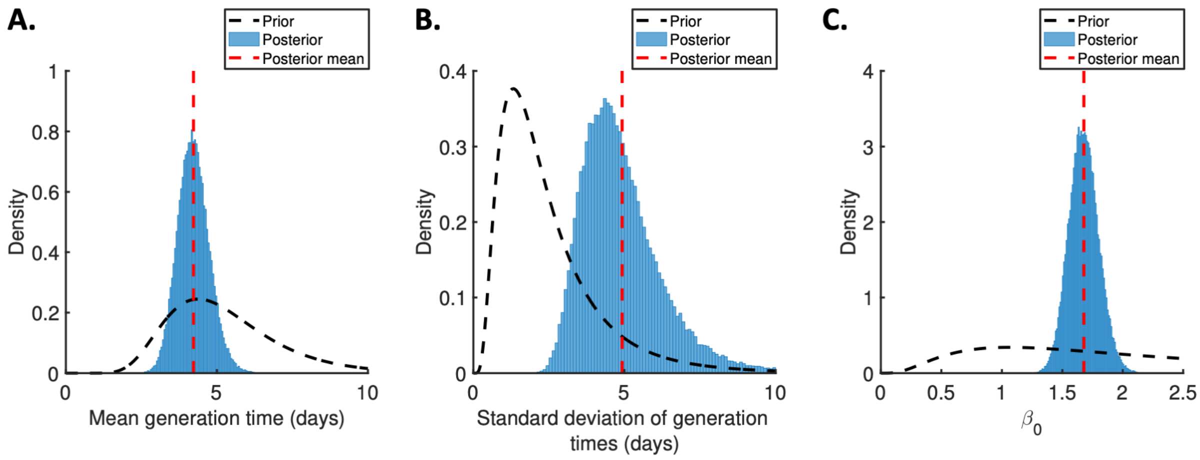


**Figure S1. Posterior distributions for the independent transmission and symptoms model.** Prior distributions (black dotted lines), posterior distributions (blue bars), and posterior means (vertical red dotted lines) for fitted parameters in the independent transmission and symptoms model. A. Mean generation time. B. Standard deviation of generation times. C. Overall infectiousness, $\beta_0$.

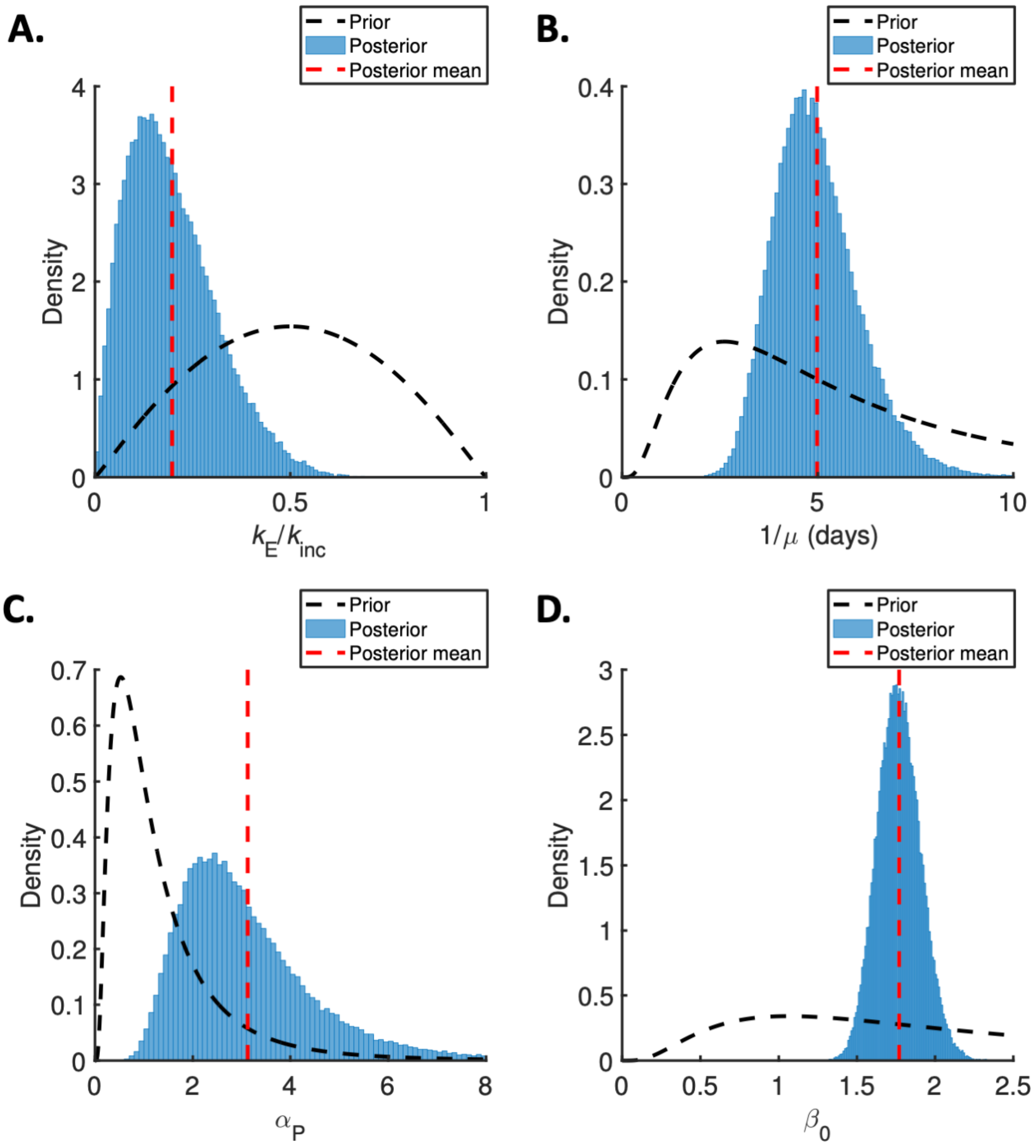


**Figure S2. Posterior distributions for the mechanistic model.** Prior distributions (black dotted lines), posterior distributions (blue bars), and posterior means (vertical red dotted lines) for fitted parameters in the mechanistic model. A. Ratio of mean durations of the latent (E) and incubation (E+P) periods, $k_E/k_{inc}$. B. Mean symptomatic infectious (I) period, $1/\mu$. C. Ratio of transmission rates in the P and I stages, $\alpha_P$. D. Overall infectiousness, $\beta_0$.

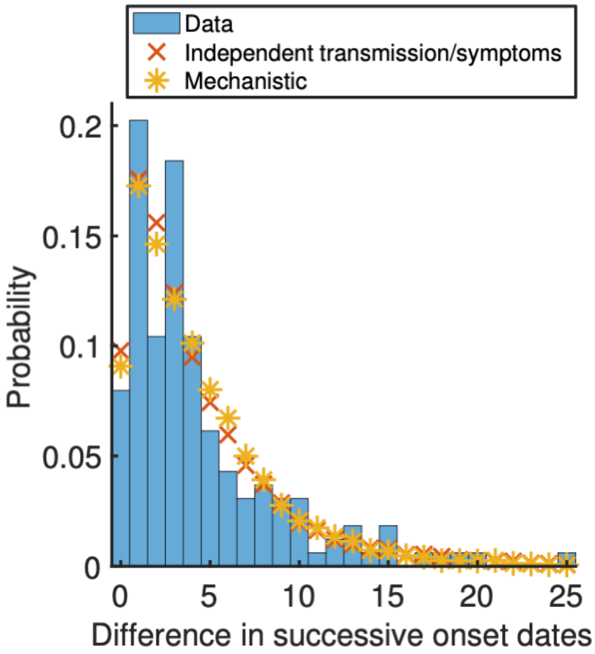


**Figure S3. Observed and model-predicted distributions of intervals between successive symptom onset dates.** Using posterior mean parameter estimates, we predicted the distribution of the difference in successive symptom onset dates (this differs from the serial interval because an infected individual may not have been infected by the previous household member to develop symptoms) under the fitted independent transmission and symptoms model (red crosses) and mechanistic model (yellow stars). These distributions were compared to the UK household data (blue bars). The distributions for the fitted models were obtained by generating synthetic data from 16,700 households using the same distribution of household sizes as in the UK data.

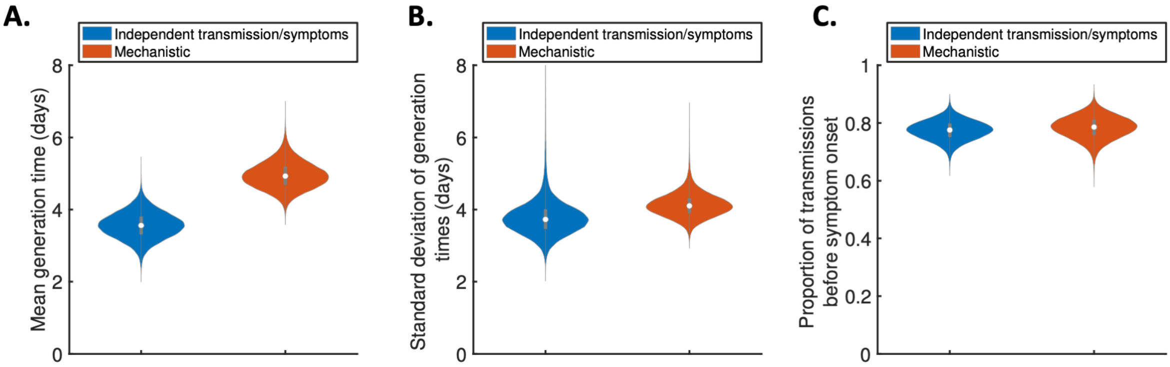


**Figure S4. Generation times within study households.** Violin plots indicating posterior distributions of the mean (A) and standard deviation (B) of realised generation times in the study households, and the proportion of transmissions occurring prior to symptom onset (C), for the independent transmission and symptoms model (blue) and mechanistic model (red).

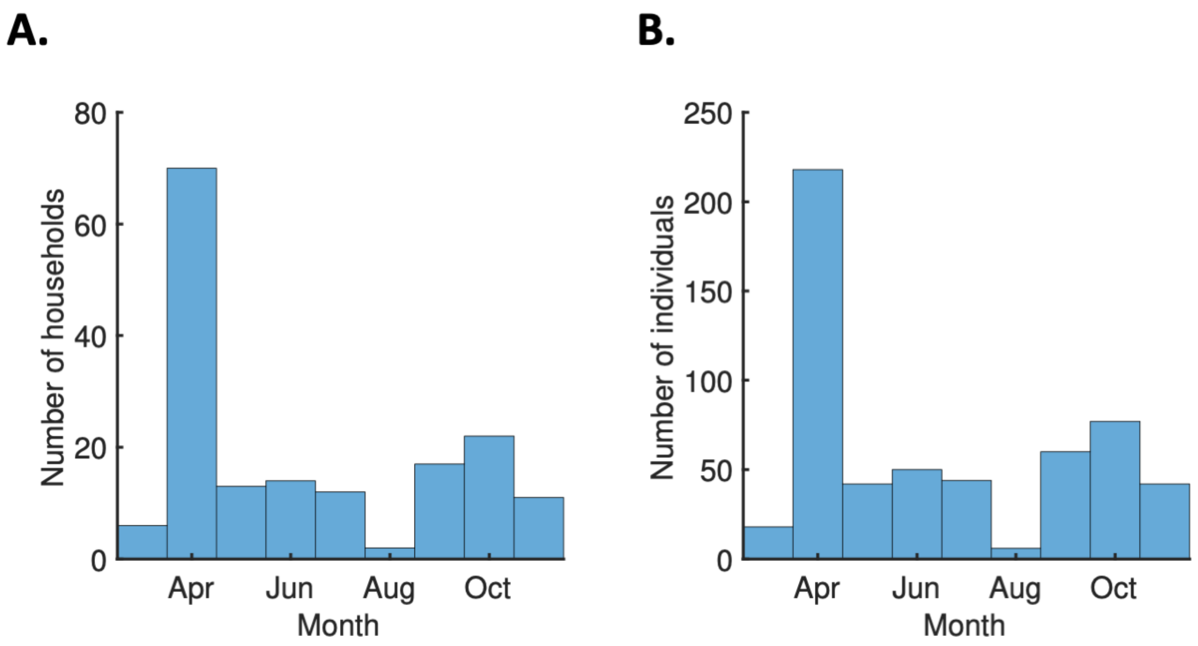


**Figure S5. Number of study households and household members by recruitment month.** A. Bars indicating the number of households recruited each month into the study from which we obtained the household transmission data used in our analyses, from March to November 2020. B. Equivalent panel showing the total number of individuals within the households recruited each month.

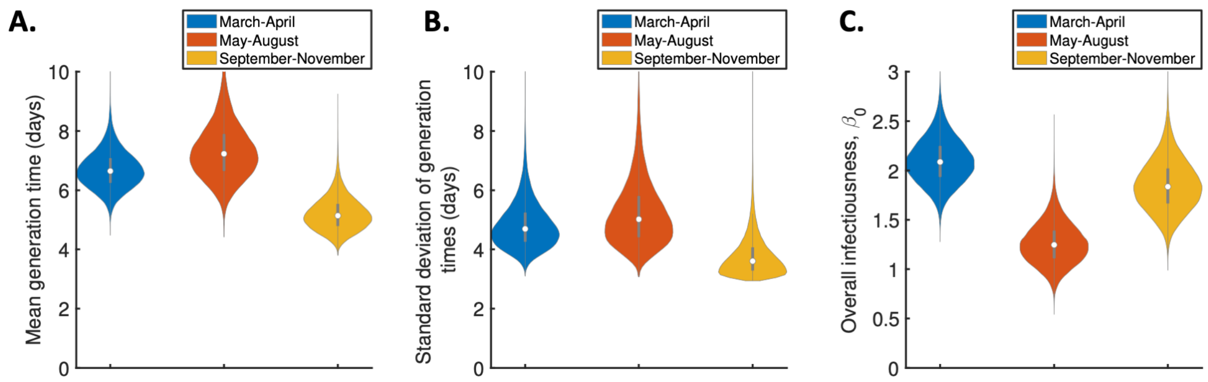


**Figure S6. Temporal changes in the generation time distribution for the mechanistic model.** Violin plots indicating posterior distributions of the mean generation time (A), standard deviation of generation times (B), and overall infectiousness parameter, $\beta_0$ (C), for the mechanistic model fitted to data from March-April (blue), May-August (red) or September-November 2020 (orange).

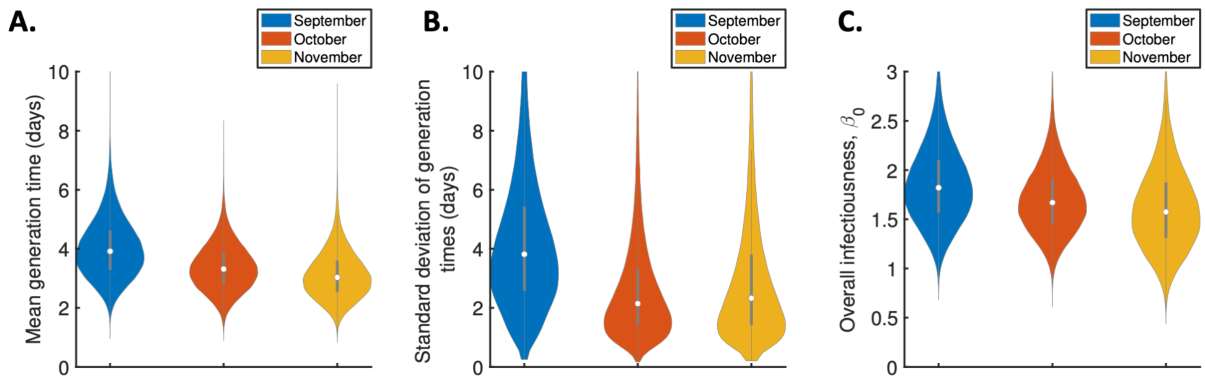


**Figure S7. Monthly changes in the generation time distribution from September-November 2020.** Violin plots indicating posterior distributions of the mean generation time (A), standard deviation of generation times (B), and overall infectiousness parameter, $\beta_0$ (C), for the independent transmission and symptoms model fitted to data from September (blue), October (red) or November 2020 (orange).

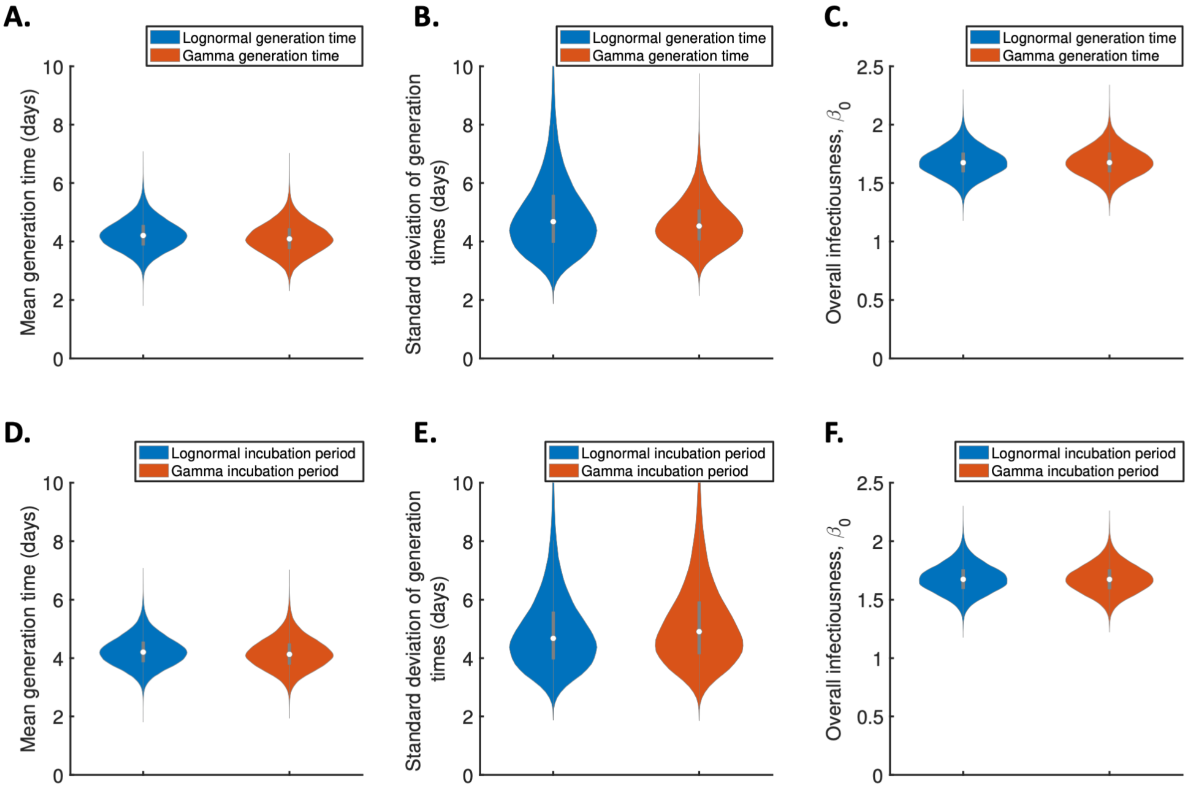


**Figure S8**. **Sensitivity of results to the functional forms of the generation time and incubation period distributions for the independent transmission and symptoms model.** A-C. Violin plots indicating posterior distributions of the mean generation time (A), standard deviation of generation times (B), and overall infectiousness parameter, $\beta_0$ (C), for the independent transmission and symptoms model, when the generation time was assumed to follow either a lognormal (blue, as in the main text) or a gamma (red) distribution, and the incubation period distribution followed a lognormal distribution (as in the main text). D-F. Equivalent panels to A-C, instead comparing cases where the incubation period was assumed to follow either a lognormal distribution (blue, as in the main text) or a gamma distribution with the same mean and standard deviation (red), and the generation time followed a lognormal distribution (as in the main text).

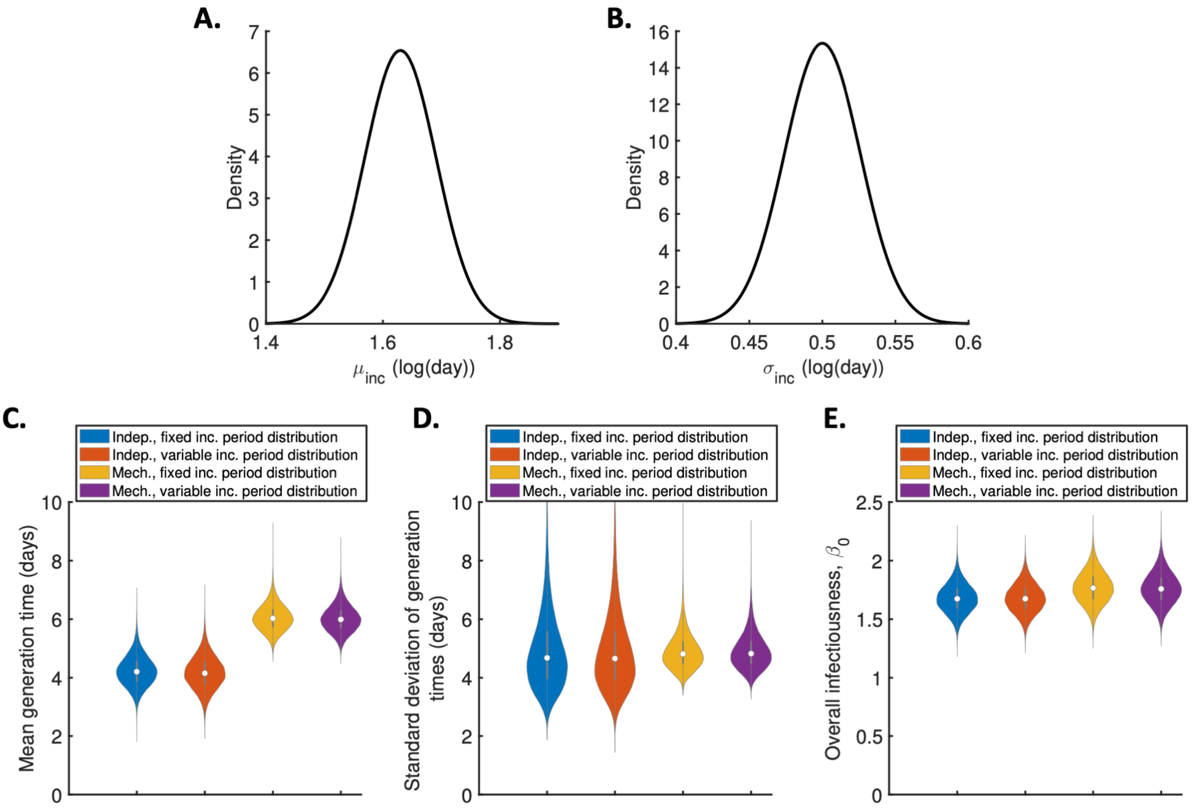


**Figure S9. Sensitivity of the results to the incubation period distribution.** A-B. Uncertainty in the incubation period distribution was accounted for by updating the parameters, $\mu_{inc}$ and $\sigma_{inc}$, of a lognormal incubation period distribution (these parameters represent the mean and standard deviation of the natural logarithm of the incubation period distribution, respectively) alongside unknown model parameters in the MCMC procedure. Independent normal prior distributions (truncated at zero) consistent with the 95% confidence intervals obtained in [2] were assumed for $\mu_{inc}$ (prior mean 1.63 log(day), standard deviation 0.061, 95% CrI 1.51-1.75; panel A) and $\sigma_{inc}$ (prior mean 0.5 log(day), standard deviation 0.026, 95% CrI 0.45-0.55; panel B). This incubation period was used directly when evaluating the likelihood in the independent transmission and symptoms model. In the mechanistic model, we assumed a gamma distributed incubation period with the same mean and standard deviation as a lognormal distribution with parameters $\mu_{inc}$ and $\sigma_{inc}$. C-E. Violin plots indicating posterior distributions of the mean generation time (C), standard deviation of generation times (D), and overall infectiousness parameter, $\beta_0$ (E), for the independent transmission and symptoms model with either a fixed (as in the main text; blue) or variable (i.e., accounting for uncertainty in $\mu_{inc}$ and $\sigma_{inc}$ as described above; red) incubation period distribution, and for the mechanistic model with a fixed (orange) or variable (purple) incubation period distribution.

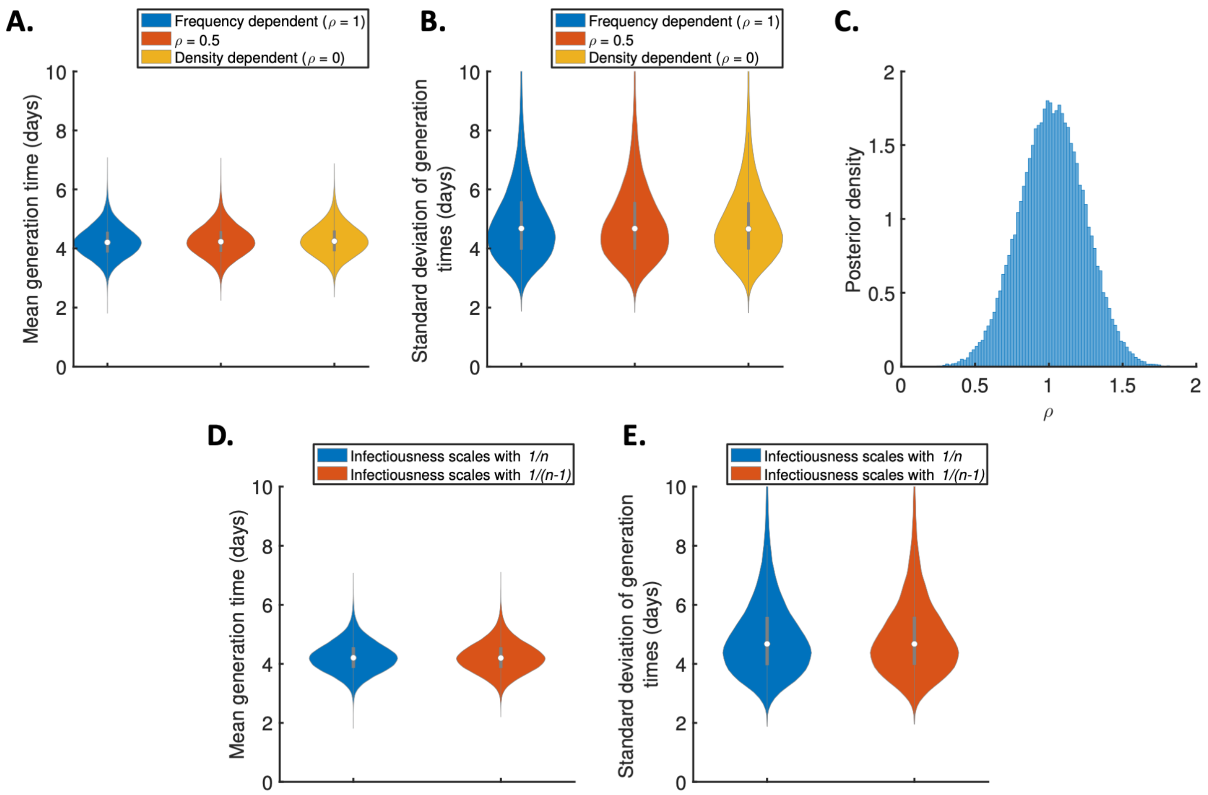


**Figure S10. Sensitivity of the results to the dependency of transmission on the household size.** A-B. Violin plots indicating the posterior distributions of the mean generation time (A) and standard deviation of generation times (B), for the independent transmission and symptoms model under different assumptions about the dependency of transmission on the household size. In these panels, infectiousness is assumed to scale with $n^{-\rho}$, where $n$ is the household size, for $\rho = 1$ (frequency-dependent transmission, blue), $\rho = 0.5$ (red), $\rho = 0$ (density-dependent transmission, orange). C. Posterior distribution of the dependency, $\rho$, when it was fitted to the data (alongside other model parameters), assuming a uniform prior for $\rho$. D-E. Equivalent panels to A-B, when infectiousness was instead assumed to scale with either $1/n$ (blue) or $1/(n-1)$ (red).

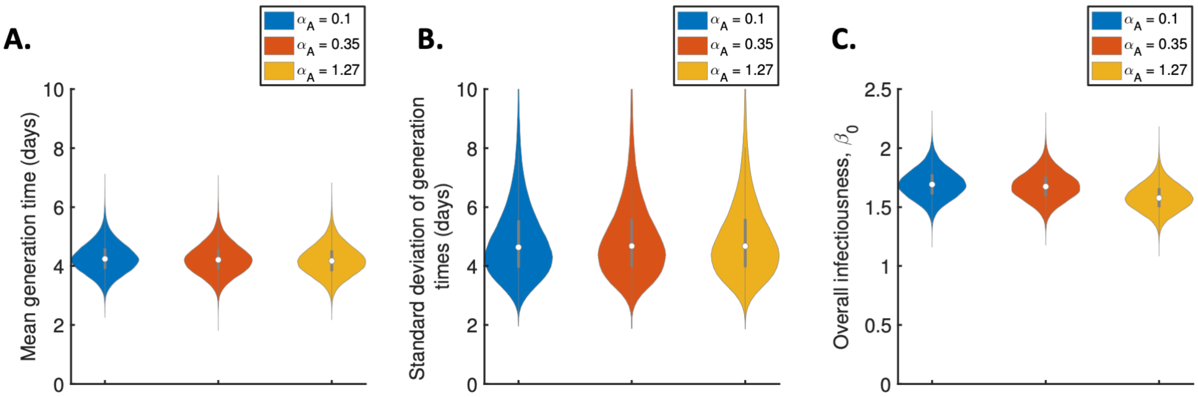


**Figure S11. Sensitivity of the results to the relative infectiousness of entirely asymptomatic infected hosts**. Violin plots indicating posterior distributions of the mean generation time (A), standard deviation of generation times (B), and overall infectiousness parameter, $\beta_0$ (C), for the independent transmission and symptoms model, when the relative infectiousness of asymptomatic hosts (compared to a host who develops symptoms, at the same time since infection) was $\alpha_A = 0.1$ (blue), $\alpha_A = 0.35$ (red) and $\alpha_A = 1.27$ (orange).

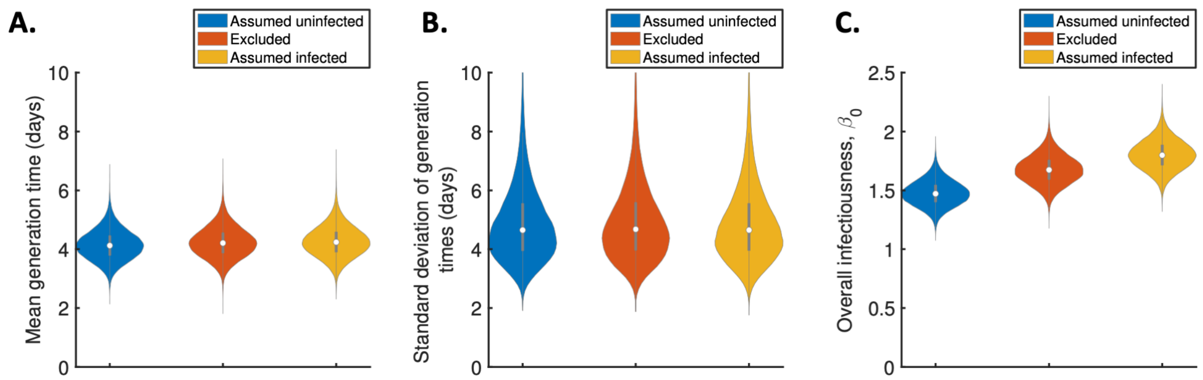


**Figure S12. Sensitivity of the results to the exclusion of hosts with unknown infection status.** A-C. Violin plots indicating the posterior distributions of the mean generation time (A), standard deviation of generation times (B), and overall infectiousness parameter, $\beta_0$ (C), for the independent transmission and symptoms model, when hosts with unknown infection status were assumed all uninfected (blue), excluded (red), or assumed all infected (orange).

297 **Supplementary Tables:**

| Parameter | Model | Interpretation | Value | Justification |
|---|---|---|---|---|
| $\alpha_A$ | Both | Relative infectiousness of entirely asymptomatic hosts | 0.35 | Taken from [3] (other values considered in sensitivity analyses) |
| Mean of natural logarithm of the incubation period | Independent transmission and symptoms | Parameter of lognormal incubation period distribution | 1.63 log(day) | Taken from [2] (uncertainty in this value considered in sensitivity analyses) |
| Standard deviation of natural logarithm of the incubation period | Independent transmission and symptoms | Parameter of lognormal incubation period distribution | 0.50 log(day) | Taken from [2] (uncertainty in this value considered in sensitivity analyses) |
| $k_{inc}$ | Mechanistic | Shape parameter of gamma incubation period distribution | 3.5 | Consistent with mean and standard deviation in [2] |
| $1/\gamma$ | Mechanistic | Mean incubation period | 5.8 days | Consistent with mean and standard deviation in [2] |
| $k_I$ | Mechanistic | Shape parameter of (gamma) symptomatic infectious period distribution | 1 | Assumed |

298

299 **Table S1.** Assumed (not fitted) parameter values used for the two models that we considered.

| Parameter | Prior | Posterior mean (95% CrI) |
|---|---|---|
| Mean generation time | Lognormal(1.6,0.35) [prior median 5.0 days, 95% CI 2.5-9.8 days] | 4.2 days (3.3-5.3 days) |
| Standard deviation of generation times | Lognormal(0.7,0.65) [prior median 2.0 days, 95% CI 0.6-7.2 days] | 4.9 days (3.0-8.3 days) |
| Overall infectiousness parameter, $\beta_0$ | Lognormal(0.7,0.8) [prior median 2.0, 95% CI 0.4-9.7] | 1.7 (1.4-1.9) |

**Table S2.** Fitted parameters in the independent transmission and symptoms model, the prior distributions used, and the posterior means and 95% credible intervals obtained.

| Parameter | Prior | Posterior mean (95% CrI) |
|---|---|---|
| Ratio of mean durations of the latent (*E*) and incubation (*E*+*P*) periods, $k_E/k_{inc}$ | Beta(2.1,2.1) [prior median 0.5, 95% CI 0.1-0.9] | 0.2 (0.03-0.5) |
| Mean symptomatic infectious (*I*) period, $1/\mu$ | Lognormal(1.6,0.8) [prior median 5.0 days, 95% CI 1.0-23.8 days] | 5.0 days (3.2-7.5 days) |
| Ratio of transmission rates in the *P* and *I* stages, $\alpha_P$ | Lognormal(0,0.8) [prior median 1.0, 95% CI 0.2-4.8] | 3.1 (1.2-6.9) |
| Overall infectiousness parameter, $\beta_0$ | Lognormal(0.7,0.8) [prior median 2.0, 95% CI 0.4-9.7] | 1.8 (1.5-2.1) |

**Table S3.** Fitted parameters in the mechanistic model, the prior distributions used, and the posterior means and 95% credible intervals obtained.
